# Cardiovascular disease burden, trends, and projections in Vietnam, 1990-2050: a first comprehensive national analysis from the Global Burden of Disease Study 2023

**DOI:** 10.64898/2026.05.13.26353134

**Authors:** Thien Tan Tri Tai Truyen, Phuc Nhan Bao Le, Bao Minh Ton Luu, Khanh Ly Le, Thi My Linh Nguyen, Ha Quoc Tin Nguyen, Kieu Anh Tho Pham, Huong-Dung Thi Nguyen

**Affiliations:** School of Medicine, Nam Can Tho University, Can Tho, Vietnam; University of Medicine and Pharmacy at Ho Chi Minh City, Ho Chi Minh City, Vietnam; Can Tho University of Medicine and Pharmacy, Can Tho, Vietnam; Pham Ngoc Thach University of Medicine, Ho Chi Minh City, Vietnam; Tay Do University, Can Tho, Vietnam

**Keywords:** Cardiovascular disease, Risk factors, Disability-adjusted life years, Mortality Forecasting, Vietnam

## Abstract

**Introduction:** Cardiovascular disease (CVD) remains Vietnam’s leading cause of mortality, yet no comprehensive national analysis of burden trends and future projections exists. This study characterizes Vietnam’s CVD burden from 1990 to 2023 and projects burden through 2050.

**Methods:** Using Global Burden of Disease 2023 data, we analyzed CVD prevalence, incidence, mortality, and disability-adjusted life years (DALYs) in Vietnam from 1990 to 2023, stratified by sex and age. Joinpoint regression quantified temporal trends. Decomposition analysis separated contributions of population growth, aging, and epidemiological change. ARIMA modeling, validated against pre-pandemic and COVID-19 periods, projected burden through 2050.

**Results:** Despite age-standardized CVD prevalence below global estimates, stroke mortality and DALYs rates exceeded global benchmarks. Age-standardized CVD mortality (ASMR) declined significantly (average annual percentage change [APC]:-1.34%), yet absolute deaths nearly doubled from 121,611 to 223,068. Population aging contributed 140.9% to observed mortality increases while epidemiological improvements averted over 102,000 deaths. Male age-standardized CVD mortality was approximately twice that of females. High systolic blood pressure remained the leading attributable risk factor, while high BMI and alcohol use showed the largest rank escalations. CVD incidence reversed its declining trend during 2019–2023 (APC:+0.69%). By 2050, ASMR are projected to decline by 51.0% (218.8 to 107.1 per 100,000 [95%CI: 64.1–150.2]), while absolute deaths are projected to increase by 43.4% (206,677 to 296,335 [95%CI: 272,323–320,348]).

**Conclusions:** Vietnam faces a demographic paradox of improving age-specific outcomes alongside a rising absolute burden driven by population aging, demanding urgent reorientation toward aging-specific prevention, hypertension control, and chronic cardiovascular care.

**Key messages:** *What is already known on this topic:* Vietnam’s cardiovascular disease burden has been characterized by stroke dominance over ischemic heart disease, ranking 4th in stroke mortality among Southeast Asian nations, with regional analyses projecting rising crude CVD mortality driven by demographic aging. However, existing evidence has been fragmented — limited to single-metric analyses, stroke-specific reports, or brief country summaries using GBD 2019 data — with no comprehensive national analysis quantifying the contributions of population growth, aging, and epidemiological change to mortality trends, nor validated projections incorporating the COVID-19 period.

*What this study adds:* This study provides the first comprehensive national analysis of Vietnam’s CVD burden using GBD 2023 data, revealing a demographic paradox wherein a 51.0% projected decline in age-standardized mortality by 2050 coexists with a 43.4% rise in absolute deaths driven overwhelmingly by population aging — which contributed 140.9% to observed mortality increases despite advances in prevention and treatment averting over 102,000 deaths. Decomposition analysis, rigorously validated ARIMA forecasting, and sex-disaggregated analyses collectively demonstrate that Vietnam’s CVD challenge is fundamentally demographic rather than clinical.

*How this study might affect research, practice or policy:* These findings demand urgent reorientation of Vietnam’s predominantly acute-care cardiovascular infrastructure toward chronic and geriatric care models, population-wide hypertension control, and targeted management of rapidly rising metabolic risks including high BMI, LDL cholesterol, and alcohol use. Sex-specific prevention strategies are needed to address behavioral risks in men and late-life cardiovascular burden in women. This evidence base provides Vietnamese and Western Pacific policymakers with the country-specific projections needed to act before the demographic transition becomes irreversible.

## Introduction

Cardiovascular disease (CVD) remains the leading cause of death worldwide.^1^ According to the Global Burden of Disease (GBD) study, global CVD deaths increased by 25–50% between 1990 and 2019, with projections suggesting this burden will reach 35–36 million deaths annually by 2050.^2^ Notably, approximately 80% of CVD deaths occur in low- and middle-income countries (LMICs), with a substantial proportion representing premature mortality among working-age adults.^3^ Vietnam exemplifies the growing cardiovascular burden in LMICs. Despite sustained efforts to control non-communicable diseases over three decades, CVD remains the leading cause of death, accounting for approximately 31% of all mortality, with stroke alone contributing 16%.^4,5^ Vietnam’s rapid economic transformation following the “Doi Moi Renovation” policies has been accompanied by significant demographic and epidemiological transitions, including population aging, urbanization, and shifts in lifestyle factors. The COVID-19 pandemic may have further exacerbated Vietnam’s cardiovascular burden. Early and strict public health measures effectively controlled viral transmission but significantly disrupted cardiovascular healthcare services, particularly for high-risk populations. Evidence indicates that cardiovascular care activities declined by 60–100% during the pandemic peak, and interruptions in chronic disease management are expected to increase CVD incidence and mortality through inadequate control of modifiable risk factors.^6,7^

Despite extensive global research on CVD burden utilizing GBD data, comprehensive analyses of long-term cardiovascular disease trends in Vietnam remain scarce, and the demographic and epidemiological risk factor factors driving the evolving CVD burden have not been systematically evaluated. Furthermore, the potential impact of health system disruptions during the COVID-19 pandemic, on CVD mortality patterns has not been assessed. To address these knowledge gaps, we analyzed data from the GBD Study 2023 with three primary objectives: (1) to characterize comprehensively the CVD burden in Vietnam in 2023 relative to Southeast Asian (SEA) and global estimates, and to analyze longitudinal trends in mortality, incidence, prevalence, and disability across 1990–2023, stratified by sex and age group; (2) to decompose the relative contributions of population growth, population aging, changes in age-specific mortality rate, and specific risk factors to the observed mortality trends; and (3) to project CVD mortality burden through 2050, providing critical evidence to inform cardiovascular disease prevention strategies and optimize healthcare resource allocation.

## Methods

Data were retrieved from the GBD 2023 database, publicly released since October 2025 (available at https://vizhub.healthdata.org/gbd-results/). The GBD 2023 database was collected and organized by the Institute for Health Metrics and Evaluation at the University of Washington. The aims and methods used in the GBD 2023 estimates have been described in detail elsewhere.^8^ For our analysis, we utilized primarily the cause of death (CoD) estimate, which provides data on the epidemiologic parameters of 292 causes of death by age group (0 to 95+ years), sex (female, male), location, and year. We extracted age-specific CVD mortality data and population estimates for Vietnam spanning from January 1, 1990, to December 31, 2023. Data were stratified into 20 age groups: <5 years, 5–9 years, 10–14 years, and continuing in 5-year increments through 90–94 years, with a final category of 95 years and older. The complete CVD aggregate encompasses eleven conditions: ischemic heart disease, stroke, hypertensive heart disease, atrial fibrillation and flutter, cardiomyopathy and myocarditis, rheumatic heart disease, non-rheumatic valvular heart disease, peripheral artery disease, aortic aneurysm, endocarditis, and other cardiovascular and circulatory diseases, each defined according to standardized GBD case definition (Supplemental Methods).^8^

### Risk factors analysis

The GBD 2023 study analyzes 88 modifiable risk factors organized into a 4-level hierarchical structure using the comparative risk assessment (CRA) framework. Level 1 represents the broadest categories, including behavioral, metabolic, and environmental and occupational risk factors, while Level 4 provides the most granular classifications. In this study, we examined the contribution of various risk factors to CVD-related deaths and disability-adjusted life years (DALYs), beginning with Level 1 categories and progressing through Levels 2, 3, and 4.^9^

For each risk–outcome pair, the population attributable fraction (PAF) was calculated, representing the proportional reduction in disease burden that would occur if population exposure to a risk factor were reduced to the theoretical minimum risk exposure level (TMREL). Risk-attributable burden was then quantified by multiplying the PAF by the total disease burden (deaths or DALYs) associated with specific cardiovascular outcomes. The detailed methodology for exposure estimation, relative risk quantification, and PAF calculation has been described elsewhere.^9^

### Decomposition Analysis

To quantify the relative contributions of demographic and epidemiological factors to changes in CVD mortality burden, we employed the Das Gupta decomposition method, a widely used technique in population health research.^10^ This approach partitions the observed change in absolute number of deaths between two time points into three distinct components: population growth, population aging, and epidemiological change (Supplemental Methods).

A negative epidemiological change indicates that age-specific mortality rates declined over the study period (improvement), while a positive value indicates increasing age-specific rates (deterioration). We calculated age-specific mortality rates as deaths per 100,000 population for each age group in 1990 and 2023. The percentage contribution of each component to the total change was computed by dividing the absolute change from each component by the total observed change in deaths and multiplying by 100.

### Projection of CVD mortality burden in Vietnam to 2050

To estimate the future CVD mortality burden in Vietnam through 2050, projection modeling was conducted separately for two metrics: age-standardized mortality rates (ASMR) and absolute death counts. The analytic period (1990–2023) was stratified into three phases: (1) a training period (1990–2009, n=20 years) for initial model fitting; (2) a pre-pandemic validation period (2010–2019, n=10 years) for out-of-sample performance assessment under stable epidemiological conditions; and (3) a COVID-19 period (2020–2023, n=4 years) for stress-testing model resilience under pandemic disruption. This three-phase strategy provides stronger evidence of model generalizability than single-split validation. The complete methodology is described in detail in the supplementary material (Supplemental Methods). In brief, we employed forecasting models previously validated in published studies, including Poisson regression, autoregressive integrated moving average (ARIMA), and exponential smoothing (ETS).^11,12^ All candidate models were fitted to the training data (1990–2009) and subsequently projected to 2010–2019, with projections compared against actual 2010–2019 data using root-mean-squared error (RMSE), mean error (ME), and Akaike Information Criterion (AIC) to evaluate model performance and bias. Lower RMSE, ME, and AIC values indicate superior predictive accuracy. This procedure identified ARIMA(0,1,2) and ARIMA(0,1,1) with drift as the optimal models for ASMR and absolute death counts, respectively (Supplemental Table S1). Final models were fitted on the complete pre-pandemic dataset (1990–2019, n=30) and used to generate 27-step-ahead forecasts from 2024 to 2050. Prediction intervals were derived from forecast standard errors under the assumption of normally distributed residuals, yielding 80% and 95% prediction intervals. Goodness of fit was assessed using the Ljung-Box test at lag 6, determined as min(10, n/5) for an annual series of length n=30. A p-value greater than 0.05 indicated that no significant autocorrelation remained in the model residuals, suggesting that the model had adequately captured the temporal patterns in the data and that the residuals behaved randomly as expected. Model performance was additionally assessed over the COVID-19 period (2020–2023) by comparing projected values against actual observed GBD 2023 estimates, evaluated using RMSE, mean absolute percentage error (MAPE), and 95% prediction interval coverage.

### Statistical Analysis

We reported a comprehensive range of epidemiological metrics, including crude numbers, percentages of deaths, and age-standardized rates (ASR) per 100,000 population for incidence, prevalence, mortality, and disability-adjusted life years (DALYs). ASRs were calculated by determining age-specific rates for each group and weighting them according to the GBD world standard population distribution to ensure comparability across different age structures and time periods. To account for potential error and modeling variance, 95% uncertainty intervals (UIs) were derived using the 2.5th and 97.5th percentiles.

Temporal trends over the 33-year period were quantified using joinpoint regression models using the Joinpoint Regression Program, Version 5.4.0 (Statistical Research and Applications Branch, National Cancer Institute). A log transformation was applied, and the optimal number of joinpoints was identified using a permutation test with 4499 permutations. For each segment (joinpoint), this analysis generated the Annual Percent Change (APC) for specific time segments and the Average Annual Percent Change (AAPC) for the overall period from 1990 to 2023 along with their 95% CI. A significant increase or decrease in prevalence, mortality, and DALYs rates was identified if *P* < .05; otherwise, the rates were considered stable. Furthermore, the study investigated the burden of heart failure (HF) as a secondary consequence of CVD, reporting age-standardized prevalence and years lived with disability (YLDs). Decomposition and projection analysis were performed using R software version 4.5.0 (R Foundation for Statistical Computing, Vienna, Austria) using the “*tidyverse*”, “*forecast*”,” *tseries*” packages, and visualizations created using “*ggplot2*” (version 3.4.0).

### Ethical Considerations

This study utilized aggregate, de-identified data from the GBD 2023 database, which is publicly available. No individual-level data were accessed, and therefore ethical approval was not required.

## Results

### The Burden of CVD in Vietnam in 2023

In 2023, Vietnam’s ASPR of CVD was 5,017.94 (95% UI: 4,742.61–5,339.70) per 100,000 population, comparable to the SEA estimate of 5,415.11 (95% UI: 5,116.00–5,794.63) but significantly lower than the global average of 6,988.81 (95% UI: 6,609.59–7,472.39) (Table 1). The ASMR of 208.36 (95% UI: 172.05–243.20) per 100,000 was comparable to both SEA (246.31, 95% UI: 220.66–269.60) and global estimates (214.94, 95% UI: 194.23–229.57). Similarly, the ASMR of 4,877.13 (95% UI: 4,121.10–5,596.20) per 100,000 was comparable to the global level (4,863.82, 95% UI: 4,460.87–5,192.31) but significantly lower than the SEA regional estimate (6,255.93, 95% UI: 5,590.94–6,801.67).

**Table 1.** Age-standardized rates of cardiovascular disease burden in Vietnam, Southeast Asia, and globally for both sexes, 2023.

|  | Vietnam | Southeast Asian Nations | Global |
| --- | --- | --- | --- |
| **Cardiovascular disease** |  |  |  |
| Prevalence | 5017.94<br>(4742.61 - 5339.70) | 5415.11<br>(5116.00 - 5794.63) | 6988.81<br>(6609.59 - 7472.39) |
| Incidence | 540.01<br>(505.35 - 581.29) | 556.55<br>(518.58 - 606.27) | 590.39<br>(555.61 - 645.46) |
| Deaths | 208.36<br>(172.05 - 243.20) | 246.31<br>(220.66 - 269.60) | 214.94<br>(194.23 - 229.57) |
| DALYs | 4877.13<br>(4121.10 - 5596.20) | 6255.93<br>(5590.94 - 6801.67) | 4863.82<br>(4460.87 - 5192.31) |
| **Stroke** |  |  |  |
| Prevalence | 1447.43<br>(1386.31 - 1510.54) | 1481.84<br>(1392.65 - 1578.86) | 1169.83<br>(1097.43 - 1256.73) |
| Incidence | 236.27<br>(216.55 - 255.38) | 219.18<br>(200.11 - 241.55) | 148.05<br>(134.67 - 165.56) |
| Deaths | 119.47<br>(94.51 - 143.62) | 118.52<br>(101.84 - 136.17) | 75.85<br>(67.64 - 83.44) |
| DALYs | 2806.25<br>(2287.77 - 3340.96) | 3030.84<br>(2633.70 - 3457.13) | 1737.13<br>(1564.30 - 1916.54) |
| **Ischemic Heart Disease** |  |  |  |
| Prevalence | 1278.65<br>(1086.56 - 1492.71) | 1640.96<br>(1427.58 - 1875.68) | 2637.35<br>(2329.69 - 2997.93) |
| Incidence | 93.70<br>(80.82 - 108.52) | 117.00<br>(101.79 - 134.30) | 140.90<br>(122.90 - 161.45) |
| Deaths | 56.00<br>(40.69 - 74.11) | 95.41<br>(80.36 - 109.44) | 99.68<br>(89.81 - 108.31) |
| DALYs | 1240.31<br>(901.31 - 1638.38) | 2357.06<br>(1983.36 - 2703.76) | 2128.58<br>(1939.96 - 2314.66) |
| **Atrial fibrillation and flutter** |  |  |  |
| Prevalence | 593.29<br>(450.31 - 754.94) | 608.92<br>(463.46 - 779.41) | 649.41<br>(510.95 - 796.42) |
| Incidence | 51.55<br>(40.07 - 65.99) | 52.97<br>(40.82 - 67.20) | 54.75<br>(43.49 - 68.81) |
| Deaths | 4.16<br>(2.72 - 5.90) | 4.01<br>(3.06 - 4.98) | 4.40<br>(3.71 - 4.95) |
| DALYs | 99.28<br>(76.96 - 130.11) | 98.84<br>(77.08 - 130.36) | 103.84<br>(84.64 - 130.27) |
| **Rheumatic Heart Disease** |  |  |  |
| Prevalence | 439.06<br>(346.79 - 535.93) | 430.14<br>(342.32 - 523.39) | 669.62<br>(535.65 - 798.85) |
| Incidence | 20.89<br>(15.89 - 26.18) | 22.11<br>(17.13 - 27.35) | 57.53<br>(47.11 - 68.30) |
| Deaths | 1.52<br>(0.86 - 2.36) | 1.83<br>(1.14 - 2.87) | 4.40<br>(2.95 - 6.26) |
| DALYs | 88.18<br>(59.07 - 125.07) | 109.27<br>(72.17 - 159.80) | 169.04<br>(118.34 - 229.67) |
| Cardiomyopathy and myocarditis |  |  |  |
| Prevalence | 41.05<br>(33.59 - 48.99) | 31.03<br>(25.29 - 36.84) | 64.67<br>(53.18 - 75.22) |
| Incidence | 11.63<br>(9.02 - 14.50) | 12.21<br>(9.55 - 15.19) | 12.46<br>(9.86 - 15.70) |
| Deaths | 2.94<br>(1.76 - 4.33) | 2.86<br>(1.92 - 3.89) | 4.58<br>(3.86 - 5.34) |
| DALYs | 76.85<br>(44.36 - 115.51) | 80.94<br>(54.22 - 109.51) | 141.48<br>(114.13 - 171.91) |

DALYs: disability-adjusted life years
All numbers were presented as numbers per 100,000 population

Among CVD subgroups, stroke was the leading cause of mortality and imposed a significantly higher burden than global estimates. Vietnam’s stroke ASPR of 1,447.43 (95% UI: 1,386.31–1,510.54) and ASMR of 119.47 (95% UI: 94.51–143.62) per 100,000 were both significantly higher than global figures (ASPR: 1,169.83, 95% UI: 1,097.43–1,256.73; ASMR: 75.85, 95% UI: 67.64–83.44), yet comparable to the SEA region. Stroke-related AS DALY rate in Vietnam (2,806.25, 95% UI: 2,287.77–3,340.96) likewise significantly exceeded the global level (1,737.13, 95% UI: 1,564.30–1,916.54) while remaining comparable to SEA (Supplemental results).

### Analysis of CVD Burden by Sex in 2023

Sex-stratified analysis revealed substantial disparities in CVD mortality burden within Vietnam. Males consistently exhibited higher age-specific death rates than females across most age groups, a pattern particularly pronounced in the 65–84 year range; however, females showed comparable or slightly higher rates at age 95 and above (Figure S1). A similar male predominance was observed for DALY rates across working-age and older groups, while age-specific prevalence and incidence rates were broadly similar between sexes, with females marginally exceeding males at the oldest age groups. Male ASMR (303.70, 95% UI: 246.82–370.39) was approximately twice that of females (143.83, 95% UI: 114.43–177.71), a pattern consistent with SEA and global trends (Table 2) (Supplemental results).

**Table 2.**
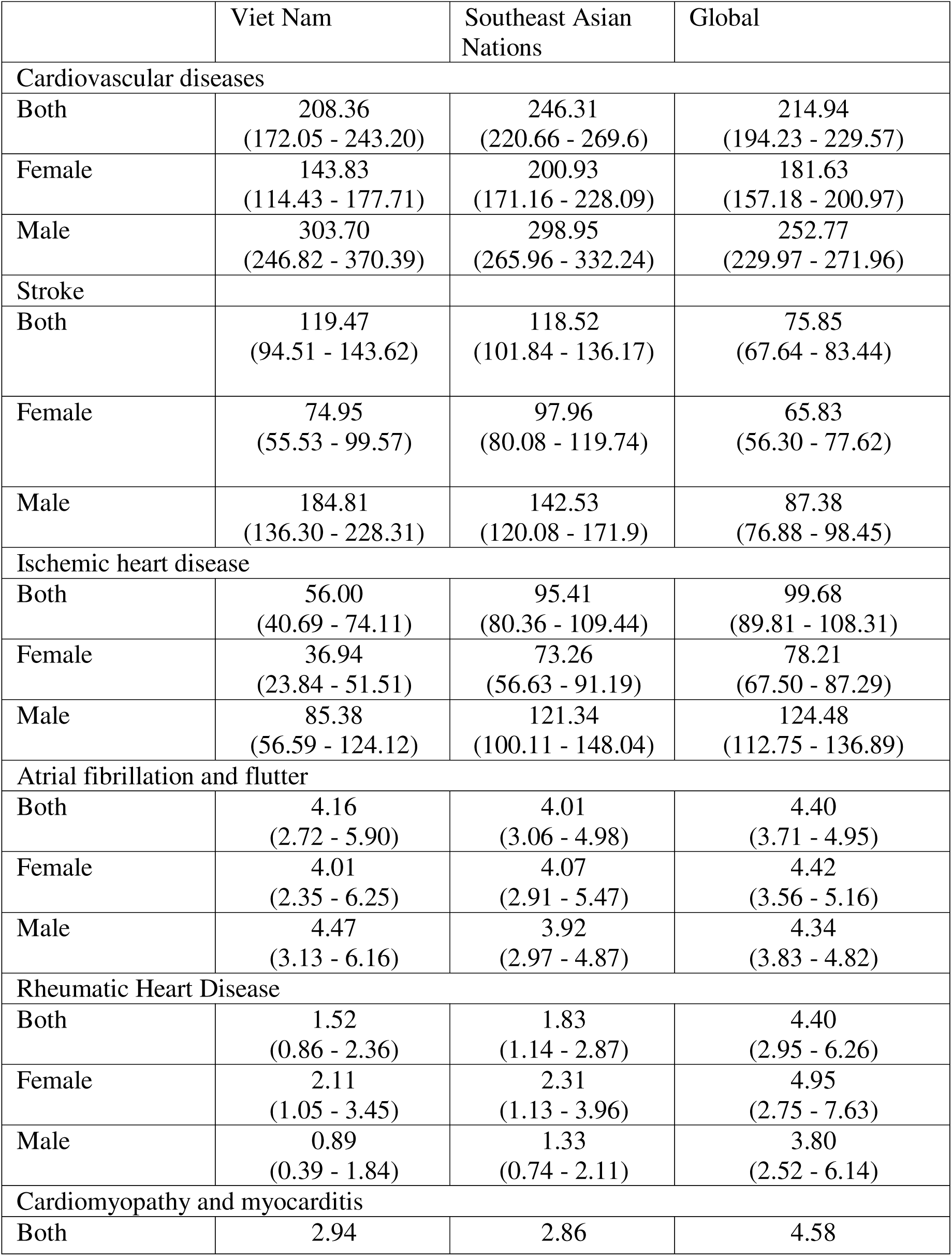
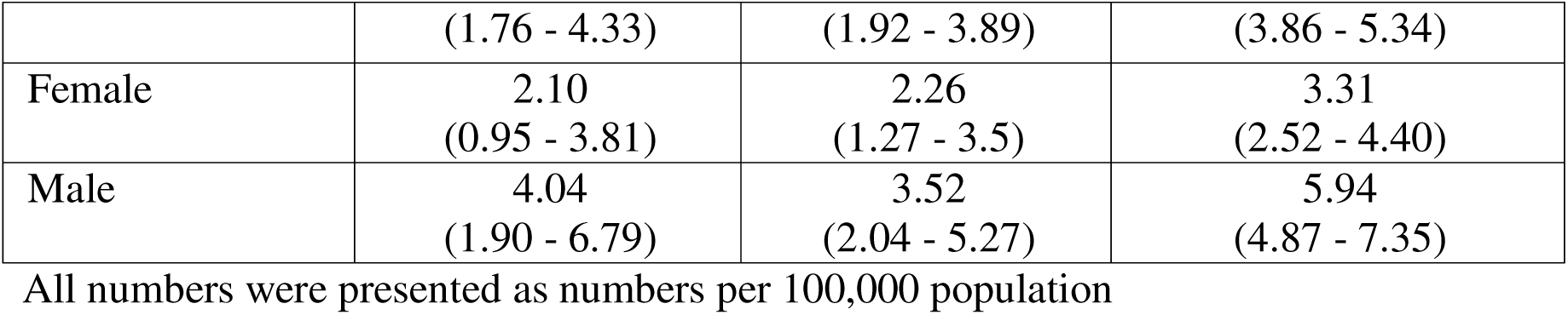
Sex-stratified age-standardized death rates (per 100,000 population) for cardiovascular disease and its subgroups in Vietnam, Southeast Asia, and globally, 2023.

### Trends in CVD Mortality Burden in Vietnam from 1990 to 2023

Between 1990 and 2023, the absolute number of CVD deaths in Vietnam increased substantially from 121,611 to 223,068, representing an 83.4% increase (101,457 additional deaths). The crude CVD mortality rate rose correspondingly from 177.4 to 216.3 per 100,000 population over the same period. Joinpoint regression analysis revealed a significant overall decline in ASMR of CVD, with an AAPC of −1.34% (95% CI: −1.41 to −1.31; p < 0.001). This improvement was significantly greater in females (AAPC: −1.59%, 95% CI: −1.63 to −1.55) than in males (AAPC: - 1.21%, 95% CI: −1.26 to −1.18) (Table S1, Figure 1). Age-standardized incidence rate similarly declined over the study period, with an overall AAPC of −0.33% (95% CI: −0.35 to −0.31; p < 0.001), with females declining significantly faster (AAPC: −0.42%, 95% CI: −0.44 to −0.41) than males (AAPC: −0.26%, 95% CI: −0.28 to −0.24). However, we identified a notable reversal from 2019 to 2023 across all groups (APC — Both: +0.69%, Female: +0.63%, Male: +0.68%), likely reflecting disruptions in CVD care attributable to the COVID-19 pandemic (Figure 1).

**Figure 1.**
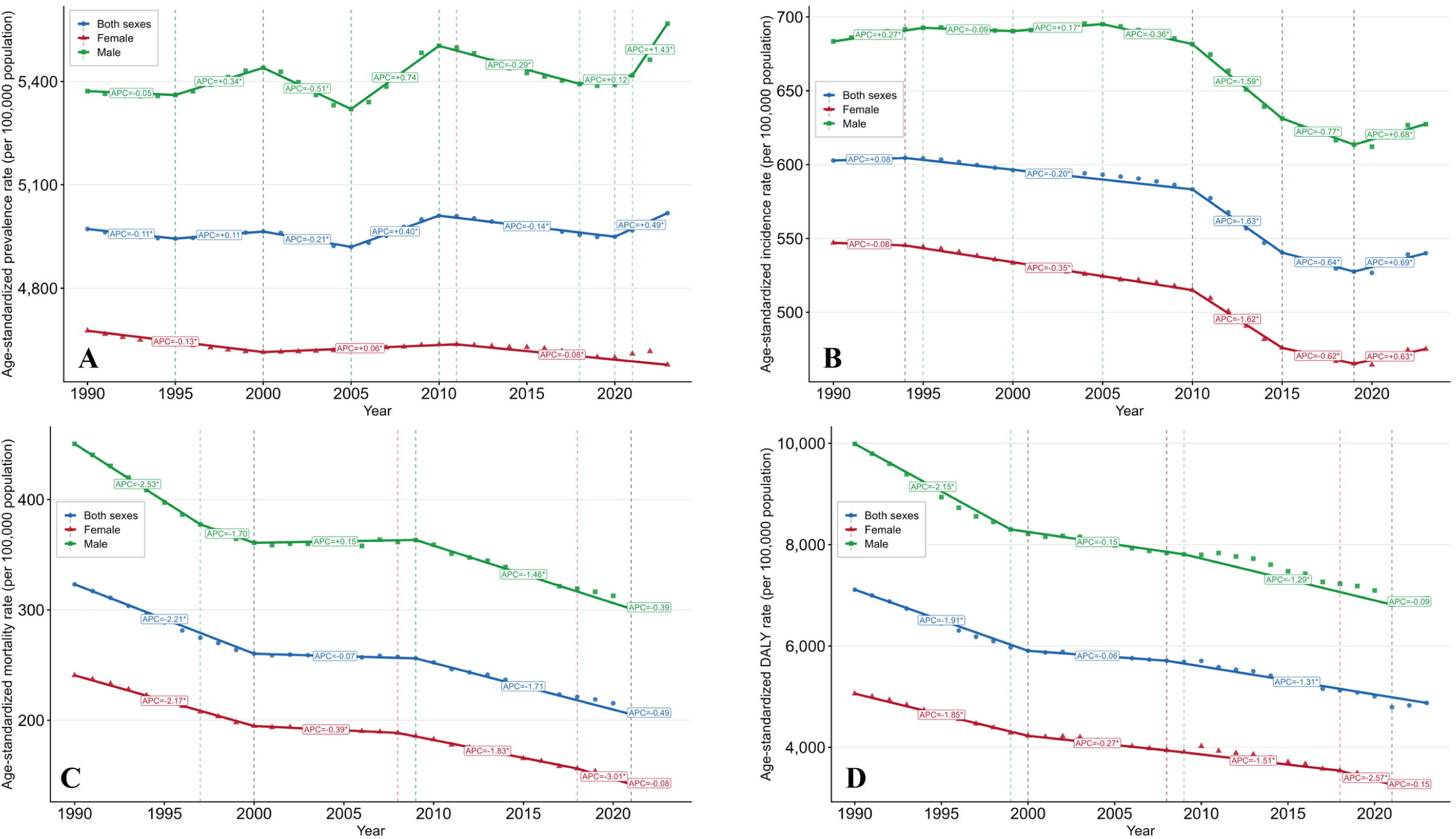
Joinpoint regression analysis of age-standardized cardiovascular burden trends in Vietnam, 1990–2023 **Figure legend.** Panels show age-standardized rates per 100,000 population for (A) prevalence, (B) incidence, (C) mortality, and (D) disability-adjusted life years (DALYs), stratified by sex. Data points represent annual observed values. Solid lines indicate fitted joinpoint regression segments, with dashed vertical lines marking estimated joinpoints (years of significant trend change). Annual percent change (APC) values are displayed for each segment; asterisks (*) denote APCs significantly different from zero (α = 0.05). Average annual percent change (AAPC) over the full study period: prevalence — both sexes +0.03%, females −0.05%, males +0.11%; incidence — both sexes −0.33%, females −0.42%, males −0.26%; mortality — both sexes −1.34%, females −1.59%, males −1.21%; DALYs — both sexes −1.19%, females −1.33%, males −1.11%. The incidence reversal observed from 2019 to 2023 (APC: both sexes +0.69%, females +0.63%, males +0.68%) is attributed to disruptions in cardiovascular care during the COVID-19 pandemic.

### Decomposition Analysis of CVD Burden Change

Decomposition analysis revealed that population aging was the dominant driver of increased CVD mortality burden over the 33-year study period (Figure S2). Population aging accounted for 142,993 additional deaths, contributing 140.9% to the total observed change. Population growth contributed to 61,409 deaths (60.5% of total change). Notably, the epidemiological change component was negative, accounting for −102,945 deaths (−101.5% of total change), indicating substantial improvements in age-specific CVD mortality rates over time. The large negative epidemiological change component demonstrates that improvements in age-specific mortality rates effectively prevented more than 102,000 CVD deaths that would have occurred if 1990 mortality rates had persisted through 2023.

### The Burden of Heart Failure related to CVD in 2023

In 2023, the ASPR of HF attributable to CVD in Vietnam was 443.55 (95% UI: 381.79–511.68) per 100,000 population. HF burden demonstrated a pronounced age gradient, with prevalence rates rising steeply across older age groups, reaching 9,580.26 and 9,963.31 per 100,000 in males and females aged 95 years and above, respectively (Figure S3). Sex-stratified analysis revealed that males tend to carry a higher ASPR (473.39, 95% UI: 407.71–549.29) and YLD rate (47.01, 95% UI: 32.03–65.31) compared to females (421.19, 95% UI: 355.57–490.74 and 40.40, 95% UI: 27.70–57.39, respectively).

### Risk Factors Associated with Death and DALYs attributed to CVD in Vietnam from 1990 to 2023

High systolic blood pressure (HSBP) was the leading attributable risk factor for both CVD deaths and DALYs in Vietnam in 1990 and 2023, contributing 199.47 (95% UI: 149.06–245.76) ASMR and 4,113.1 (95% UI: 2,997.0–5,145.6) DALYs in 1990, declining to 131.64 (95% UI: 101.68–167.53) deaths per 100,000 and 2,907.5 (95% UI: 2,277.7–3,576.9) DALYs in 2023 (Figure 2, Figure S4). Environmental and behavioral risks closely followed, with particulate matter pollution and smoking consistently ranking second and third for both deaths and DALYs throughout the study period. Notable shifts emerged among metabolic risk factors: high LDL cholesterol advanced from 5th to 4th, high BMI climbed sharply from 21st to 14th for deaths (19th to 12th for DALYs), and high fasting plasma glucose rose from 13th to 12th. High alcohol use showed one of the largest rank changes overall, rising from 26th to 15th for deaths. Conversely, dietary risks declined in ranking, including diet high in sodium (4th to 6th) and diet low in vegetables (10th to 13th for deaths).

**Figure 2.**
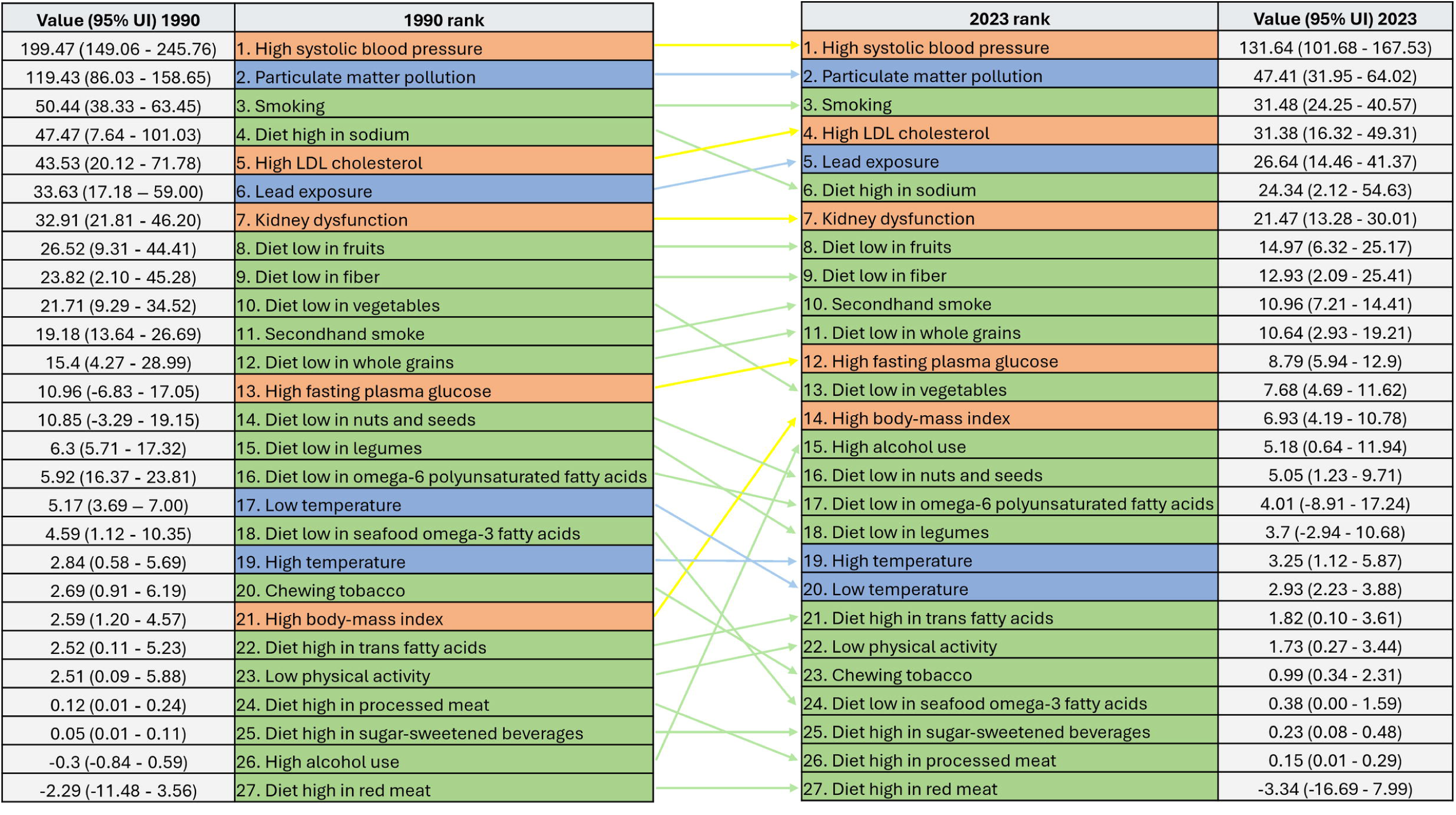
Age-standardized CVD mortality rates attributable to cardiovascular risk factors in Vietnam, 1990 and 2023 **Figure legend.** The rankings of 27 modifiable cardiovascular risk factors were displayed by their attributable age-standardized CVD mortality rate (per 100,000 population) in 1990 (left) and 2023 (right), with corresponding values and 95% uncertainty intervals (UI) shown at each end. Connecting lines illustrate rank changes between the two time points; yellow lines indicate notable upward rank shifts in metabolic risk factors (high LDL cholesterol, high body-mass index, high fasting plasma glucose) and high alcohol use, while grey lines indicate stable or downward shifts. Row colors denote risk factor categories: orange = metabolic risks, blue = environmental risks, green = dietary and behavioral risks. All estimates are derived from the Global Burden of Disease Study 2023 using the comparative risk assessment framework. CVD = cardiovascular disease; LDL = low-density lipoprotein; UI = uncertainty interval.

### Projection of CVD Burden in Vietnam to 2050

Following systematic evaluation of nine candidate models, ARIMA(0,1,2) with drift demonstrated superior performance for age-standardized death rates during the 2010–2019 validation period (RMSE: 3.40 per 100,000; MAPE: 1.38%), outperforming the second-ranked Poisson linear model by 54% (RMSE: 7.31). For absolute deaths, ARIMA(0,1,1) with drift achieved the best performance (RMSE: 1,992 deaths; MAPE: 0.92%), representing a 39% improvement over Poisson linear regression (RMSE: 3,245) (Figure S5A-B, Table S2).

Age-standardized CVD mortality rates are projected to decline by 51.0% from 218.8 per 100,000 in 2019 to 107.1 (95% CI: 64.1–150.2) in 2050, reflecting continued improvements in CVD prevention and treatment. Intermediate projections show steady decline: 179.7 (95% CI: 154.9–204.5) per 100,000 in 2030 and 143.4 (95% CI: 108.3–178.6) in 2040 (Figure 3A). Despite these declining rates, absolute CVD deaths are projected to increase by 43.4% from 206,677 in 2019 to 296,335 (95% CI: 272,323–320,348) in 2050, with intermediate projections of 238,340 (95% CI: 224,300–252,379) deaths by 2030 and 267,337 (95% CI: 247,669–287,006) by 2040 (Figure 5B).

**Figure 3.**
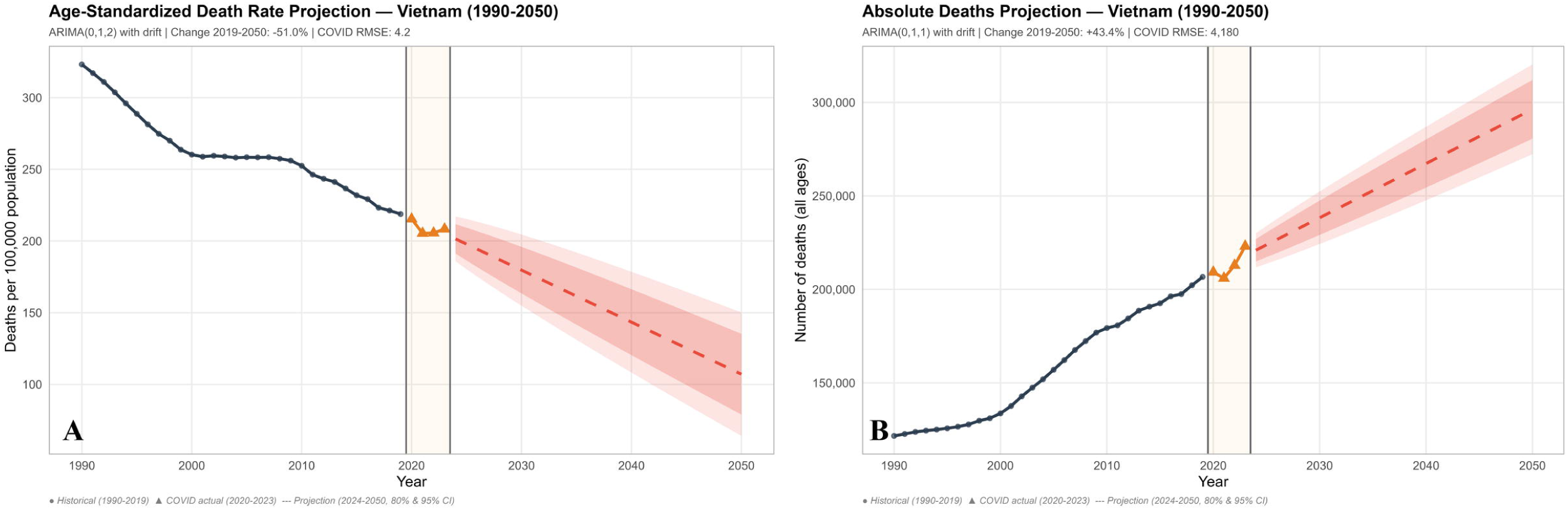
ARIMA-based projections of cardiovascular disease mortality burden in Vietnam, 1990–2050 **Figure legend.** Panel (A) shows age-standardized CVD mortality rates (deaths per 100,000 population) and Panel (B) shows absolute CVD death counts from 1990 to 2050. Navy circles represent historical observed values (1990–2019). Orange triangles indicate actual values during the COVID-19 period (2020–2023), with the shaded beige band demarcating this validation window. Dashed red lines represent projected estimates from 2024 to 2050, with dark and light red shading indicating 80% and 95% prediction intervals, respectively. Panel (A) was generated using ARIMA(0,1,2) with drift and Panel (B) using ARIMA(0,1,1) with drift, both selected based on superior out-of-sample performance over eight alternative forecasting models during the pre-pandemic validation period (2010–2019). Age-standardized mortality rates are projected to decline by 51.0% from 218.8 per 100,000 in 2019 to 107.1 (95% CI: 64.1–150.2) by 2050, while absolute deaths are projected to increase by 43.4% from 206,677 to 296,335 (95% CI: 272,323–320,348) over the same period, reflecting the divergent effects of improving age-specific mortality rates and population aging. COVID-19 validation period RMSE: 4.15 per 100,000 for age-standardized rates (coverage: 4/4 within 95% CI); 4,180 deaths for absolute counts (coverage: 3/4 within 95% CI). CVD = cardiovascular disease; ARIMA = autoregressive integrated moving average; CI = confidence interval; RMSE = root mean square error.

## Discussion

Recent regional analyses project CVD as the dominant leading cause of death across Southeast Asia through 2050, with high BMI and elevated fasting plasma glucose as the fastest-growing risk factors. Vietnam has been specifically identified as country where stroke mortality exceeds IHD, reflecting a regionally distinct risk profile.^13^ Our study provides the first comprehensive national analysis of CVD burden in Vietnam using GBD 2023 data (1990–2023, projected to 2050). Despite overall burden remaining comparable to or below regional benchmarks, Vietnam carries a disproportionately high stroke burden with ASMR and DALY rates exceeding global figures, while IHD remains comparatively lower. The divergence between declining ASMR and rising absolute deaths reflects demographic aging as the dominant driver, confirmed by decomposition analysis. Projections to 2050 forecast a 51.0% decline in age-standardized rates alongside a 43.4% rise in absolute deaths, underscoring the need to prioritize aging-specific prevention and management of rapidly rising metabolic risks, particularly high BMI and elevated LDL cholesterol.

Vietnam’s CVD profile in 2023 reflects the classic LMIC transition pattern of stroke dominance over IHD, with stroke ASMR and DALY rates significantly exceeding global benchmarks despite sustained improvements in age-specific mortality.^14,15^ This elevated stroke burden is partly driven by a higher proportion of hemorrhagic stroke relative to HICs — a subtype strongly associated with uncontrolled hypertension and carrying substantially higher case fatality than ischemic stroke.^16^ Vietnam’s 2015 national STEPS survey revealed that only 43.1% of hypertensive individuals were aware of their condition, and just 13.6% had their blood pressure managed at a health facility; at the regional level, recent study in 2019 found only 33.2% of hypertensive individuals were receiving treatment and a mere 12.2% had achieved blood pressure control — a compounding cascade of missed opportunities that directly induce hemorrhagic stroke risk at the population level.^17,18^

A central finding of this study is the demographic paradox underlying Vietnam’s CVD burden: while ASMR declined significantly over the study period, absolute CVD deaths nearly doubled from 121,611 in 1990 to 223,068 in 2023. Decomposition analysis reveals that this divergence is driven overwhelmingly by population aging. Advances in prevention and treatment effectively averted over 102,000 deaths across the study period, yet the sheer pace of Vietnam’s demographic aging exceeded what clinical improvements alone could address. This pattern is consistent with projections for the broader ASEAN region, where demographic shifts have been identified as the primary driver of rising absolute CVD burden despite improving age-standardized rates.^14^ The implication is clear: sustaining clinical progress alone will be insufficient — Vietnam urgently needs aging-specific cardiovascular prevention strategies to avoid being overwhelmed by its own demographic transition.

Our analysis reveals a dual epidemiological pattern in Vietnam’s CVD risk factor profile between 1990 and 2023: persistent dominance of traditional risk factors alongside accelerating metabolic and behavioral risks — consistent with rapid socioeconomic transformation following the “Doi Moi Renovation”. HSBP remained the single largest attributable risk factor throughout, aligning with the broader ASEAN region where it ranked first across all ten member nations, likely sustained by inadequate hypertension control.^13^ Among metabolic risks, high BMI showed the most striking escalation — rising from 21st to 14th for deaths — consistent with GBD 2021 data identifying it as the fastest-growing CVD risk factor across ASEAN. High fasting plasma glucose and LDL cholesterol similarly advanced, signaling an emerging cardiometabolic phenotype aligned with documented increases in diabetes and dyslipidemia among Vietnamese adults.^19,20^ Notably, high alcohol use showed one of the largest rank shifts reflecting Vietnam’s substantial increase in per-capita alcohol consumption.^21^ Therefore, Vietnam’s CVD prevention framework must therefore simultaneously address persistent traditional risks and rapidly emerging threats through evidence-based interventions including expanded hypertension management, obesity prevention, diabetes and dyslipidemia screening and treatment, and alcohol taxation.^22,23^

The COVID-19 pandemic introduced a measurable disruption to Vietnam’s CVD burden trajectory. Following sustained declines, age-standardized CVD incidence reversed course (APC: +0.69%), likely reflecting interruptions in cardiovascular care access, reduced health-seeking behavior, and deferred management of modifiable risk factors during the pandemic period. While CVD mortality remained largely on its projected trajectory — with ARIMA projections achieving 100% coverage during the COVID-19 validation period — the 2021 absolute death count fell slightly outside prediction bounds, suggesting transient pandemic-related mortality fluctuations. These findings highlight the vulnerability of cardiovascular care continuity to health system shocks and underscore the importance of resilient primary care infrastructure.

Projections to 2050 reinforce the important demographic paradox of this study. Despite a projected 51.0% decline in ASMR to 107.1 per 100,000 by 2050 — reflecting the “price of success” of improving prevention and treatment in a rapidly developing nation — absolute CVD deaths are projected to rise 43.4%, approaching 300,000 annually.^14^ This trajectory mirrors regional forecasts for Southeast Asia, where crude cardiovascular mortality is anticipated to rise 81.6% by 2050 despite falling age-standardized rates, driven by the rapidly aging population.

Clinically, this will manifest as an expanding burden of chronic cardiovascular sequelae — particularly heart failure,^24^ where prevalence already exceeds 9,500 per 100,000 in the oldest age cohort. Meeting this challenge will require Vietnam to reorient its predominantly acute-care cardiovascular infrastructure toward chronic and geriatric care models, ensure equitable access to evidence-based cardioprotective therapies,^25^ and fortify national health insurance to sustain coverage for a growing elderly population.

### Limitations and strengths

Several limitations warrant acknowledgment. GBD estimates rely on modelled data with wide UIs for several metrics, and Vietnam-specific inputs are relatively sparse, potentially masking urban-rural and regional heterogeneity. Risk factor rank changes reflect attributable burden rather than causality, and ARIMA projections cannot fully account for future policy interventions or climate-related shifts. Nonetheless, this study offers important strengths. It represents the first comprehensive national CVD analysis using GBD 2023 data, incorporating the COVID-19 period. The multi-metric framework — spanning prevalence, incidence, mortality, DALYs, heart failure burden, risk factors, and sex-stratified trends — provides exceptional analytical depth, while decomposition analysis adds mechanistic insight into demographic drivers absent from comparable regional analyses.

## Conclusion

Vietnam’s CVD burden between 1990 and 2023 reflects a nation at a critical epidemiological crossroads — achieving meaningful reductions in age-specific mortality while confronting an absolute burden that clinical progress alone cannot contain. The country’s stroke-dominant profile, substantial sex disparities, and rapidly shifting metabolic risk landscape collectively define a CVD fingerprint demanding targeted, context-specific responses. Projections to 2050 make the urgency unmistakable: without a fundamental reorientation toward aging-specific prevention and chronic cardiovascular care, Vietnam risks being overwhelmed by the demographic tide of its own success. These findings provide the most comprehensive national evidence base to date for Vietnamese policymakers and the broader ASEAN cardiovascular community to act decisively — and equitably — before that window closes.

## Data Availability

All data produced are available online at https://vizhub.healthdata.org/gbd-results/

## Authors contribution

Thien Tan Tri Tai Truyen: conceptualization, formal analysis, writing – original draft, writing – review & editing; Phuc Nhan Bao Le: methodology, data curation, formal analysis, visualization; Bao Minh Ton Luu: writing – original draft; Khanh Ly Le: writing – original draft, Thi My Linh Nguyen: data curation, formal analysis, writing – original draft, Ha Quoc Tin Nguyen: writing – review & editing, Kieu Anh Tho Pham: writing – review & editing, validation; Huong-Dung Thi Nguyen: conceptualization, methodology, project administration, supervision, validation, writing – review & editing

## Funding

This research received no specific grant from any funding agency in the public, commercial or not-for-profit sectors.

## Data sharing statement

All data and codes used in analysis and projections are available upon request to the corresponding author.

## Declaration of interests

The authors declare that they have no known competing financial interests or personal relationships that could have appeared to influence the work reported in this paper.

## Acknowledgements

We express our deepest thanks and appreciation to Nam Can Tho University and Can Tho University of Medicine and Pharmacy for the support from during the study period.

## Patient and Public Involvement

Patients or the public WERE NOT involved in the design, or conduct, or reporting, or dissemination plans of our research

## Notes

### Competing Interest Statement

The authors have declared no competing interest.

### Author Declarations

This study utilized aggregate, de-identified data from the GBD 2023 database, which is publicly available. No individual-level data were accessed, and therefore ethical approval was not required. https://vizhub.healthdata.org/gbd-results/)

