## Supplemental Table for "Cardiovascular disease burden, trends, and projections in Vietnam, 1990-2050: a first comprehensive national analysis from the Global Burden of Disease Study 2023"

**Supplemental materials**

### Supplemental Methods

**GBD data source**

GBD 2023 employs standardized disease definitions and classifications based on the International Classification of Diseases, Tenth Revision (ICD-10) codes. Estimates are derived through systematic review of all available data sources and advanced statistical modeling techniques, including Cause of Death Ensemble models (CODEm) and spatiotemporal Gaussian process regression (ST-GPR), to generate internally consistent estimates of mortality by cause, age, sex, and location.^1^ For Vietnam, GBD estimates incorporate data from vital registration systems, verbal autopsy studies, surveillance systems, and statistical modeling to provide comprehensive and comparable disease burden estimates across the 33-year study period (1990-2023).

**Decomposition analysis**

The decomposition was calculated as follows:

Population Growth Effect (PGE): This component represents the change in deaths attributable solely to changes in population size, assuming constant age structure and age-specific mortality rates from the baseline year (1990). It was calculated by applying the population growth factor to baseline deaths:

$$PGE=D1990 x \left( \frac{P2023}{P1990} \right)-D1990$$

where D_1990_ represents total deaths in 1990, and P_1990_ and P_2023_ represent total population in 1990 and 2023, respectively.

Population Aging Effect (PAE): This component captures the change in deaths due to shifts in age structure (population aging), holding population size and age-specific mortality rates constant at 1990 levels. It was computed as:

$$\mathrm{PAE}=\sum_{i=1}^{20} (ri1990x \frac{ni2023}{100000})-\sum_{i=1}^{20} (ri1990x \frac{ni1990}{100000})-PGE$$

where r*i*_1990_ is the age-specific mortality rate (per 100,000) for age group *i* in 1990, and n*i*_1990_ and n*i*_2023_ are the populations in age group *i* in 1990 and 2023, respectively.

Epidemiological Change Effect (ECE): This component reflects changes in age-specific mortality rates over time, representing improvements or deteriorations in disease risk, prevention, and treatment. It was calculated as the residual:

ECE = ΔD_total_ – PGE - PAE

where ΔD_total_ is the total observed change in deaths between 1990 and 2023.

**Projection Analysis**

*Stationarity Assessment*

Prior to candidate model identification, stationarity of each time series was formally assessed using the Kwiatkowski-Phillips-Schmidt-Shin (KPSS) test, which assumes stationarity as the null hypothesis — making it more conservative and better powered for short annual series of length n=30. The KPSS test was applied in trend-stationary form to the original series and in level-stationary form to the first-differenced series. For age-standardized mortality rates, the original series was non-stationary (KPSS p=0.029) but became stationary after first differencing (KPSS p=0.090), confirming d=1. Similarly, the absolute death count series was non-stationary in levels (KPSS p=0.022) but stationary after first differencing (KPSS p=0.065), confirming d=1 for both metrics.

*Candidate Model Identification via Box-Jenkins Approach^2^*

Candidate autoregressive (p) and moving average (q) orders were identified following the Box-Jenkins methodology through visual inspection of the autocorrelation function (ACF) and partial autocorrelation function (PACF) of the first-differenced series. The ACF quantifies the linear correlation between observations at varying time lags, while the PACF isolates the direct correlation at each lag after removing the influence of all intermediate lags. Together, these functions provide empirical guidance for identifying the appropriate ARIMA order:

- A PACF that cuts off sharply after lag p with a gradually decaying ACF suggests an AR(p) process
- An ACF that cuts off sharply after lag q with a gradually decaying PACF suggests an MA(q) process
- Both tailing off gradually suggests a mixed ARMA process

For both the ASMR and absolute death series, the PACF exhibited a significant spike at lag 1 followed by a sharp cutoff, indicating p ∈ [0, 1]. The ACF showed significant correlations at lags 1–3 with gradual subsequent decay, indicating q ∈ [1, 2]. ARIMA(0,1,0) — representing a pure random walk with no autocorrelation structure — was excluded as statistically uninformative. This inspection yielded five theoretically-informed ARIMA candidates: ARIMA(0,1,1), ARIMA(0,1,2), ARIMA(1,1,0), ARIMA(1,1,1), and ARIMA(1,1,2).

*Benchmark Models^3^*

Four additional benchmark models were included to ensure comprehensive comparison:

- **Auto ARIMA**: data-driven order selection via iterative AIC minimization across all feasible (p,d,q) combinations; selected ARIMA(0,2,0) for ASMR and ARIMA(0,2,1) for absolute deaths
- **Exponential Smoothing (ETS)**: state-space model with error, trend, and seasonal components automatically selected; identified as ETS(A,A,N) for both metrics — additive error, additive trend, no seasonality, appropriate for annual non-seasonal data
- **Poisson regression (linear trend)**: log-linear model assuming the outcome follows a Poisson distribution with a linear year effect: log(λ) = β₀ + β₁ × year
- **Poisson regression (quadratic trend)**: log(λ) = β₀ + β₁ × year + β₂ × year²

*ARIMA Model Specification*

The general ARIMA(p,d,q) model with drift is expressed as:

$$\phi\left( B \right)\left( 1-B \right)^{d}y_{t}= \mu+ \theta\left( B \right)\epsilon_{t}$$

where B is the backshift operator, φ(B) = 1 − φ₁B − ... − φₚBᵖ is the autoregressive polynomial, θ(B) = 1 + θ₁B + ... + θqBq is the moving average polynomial, μ is the drift constant representing the average change per period, d is the differencing order, and ε_t ~ N(0, σ²) are white noise residuals. For the selected models:

- **ARIMA(0,1,2) for ASMR**: $\left( 1-B \right)y_{t}= \mu+ \left( 1 + \theta_{1}B + \theta_{2}B^{2} \right)\epsilon_{t}$, with estimated coefficients θ₁=0.727, θ₂=0.316, drift=−3.629 per 100,000 per year
- **ARIMA(0,1,1) for absolute deaths**: $\left( 1-B \right)y_{t}= \mu+ \left( 1 + \theta_{1}B \right)\epsilon_{t}$, with estimated coefficients θ₁=0.605, drift=+2,900 deaths per year.

### Supplemental Results

*The Burden of CVD in Vietnam in 2023*

IHD ranked second in CVD mortality, with an ASPR of 1,278.65 (95% UI: 1,086.56–1,492.71) comparable to SEA (1,640.96, 95% UI: 1,427.58–1,875.68), but significantly lower than the global average (2,637.35, 95% UI: 2,329.69–2,997.93); its ASMR of 56.00 (95% UI: 40.69–74.11) was significantly lower than both SEA (95.41, 95% UI: 80.36–109.44) and global estimates (99.68, 95% UI: 89.81–108.31). AF and flutter (ASMR: 4.16, 95% UI: 2.72–5.90) and cardiomyopathy and myocarditis (ASMR: 2.94, 95% UI: 1.76–4.33) showed mortality rates comparable to both SEA and global levels, while RHD mortality (ASMR: 1.52, 95% UI: 0.86–2.36) was comparable to SEA (1.83, 95% UI: 1.14–2.87) but significantly lower than the global estimate (4.40, 95% UI: 2.95–6.26).

*Analysis of CVD Burden by Sex in 2023*

The male predominance was most striking for stroke, where male ASMR (184.81, 95% UI: 136.30–228.31) was more than double that of females (74.95, 95% UI: 55.53–99.57), with male rates significantly exceeding global estimates (87.38, 95% UI: 76.88–98.45) while female rates remained comparable across all settings. For IHD, males similarly bore a heavier burden (85.38, 95% UI: 56.59–124.12) than females (36.94, 95% UI: 23.84–51.51), with female IHD ASMR being significantly lower than both SEA (73.26, 95% UI: 56.63–91.19) and global estimates (78.21, 95% UI: 67.50–87.29). In contrast, sex differences were less pronounced for AF (male: 4.47, 95% UI: 3.13–6.16; female: 4.01, 95% UI: 2.35–6.25) and cardiomyopathy and myocarditis (male: 4.04, 95% UI: 1.90–6.79; female: 2.10, 95% UI: 0.95–3.81). Notably, RHD showed a reversed pattern, with females tend to experience higher mortality (2.11, 95% UI: 1.05–3.45) than males (0.89, 95% UI: 0.39–1.84).

*Age-Specific Mortality Rate Patterns*

Analysis of age-specific mortality rates revealed marked improvements across most age groups between 1990 and 2023 (Figure S1). For individuals aged 95 years and older, mortality rates declined from 9,327 per 100,000 in 1990 to 8,314 per 100,000 in 2023, a 10.9% reduction. Similarly, the 90-94 age group experienced a decline from 7,349 to 6,997 per 100,000 (4.8% reduction), and the 85-89 age group saw rates decrease from 5,683 to 4,557 per 100,000. Middle-aged populations (45-64 years) demonstrated particularly pronounced improvements, with mortality rates declining by 15-25% across age.

CVD mortality rates exhibited a steep age gradient throughout the study period, with rates increasing exponentially among individuals aged 60 years and older. The 95+ age group experienced mortality rates approximately 90-fold higher than the 40-44 age group in both 1990 and 2023. Notably, CVD mortality remained extremely low in individuals under 40 years, with rates below 50 per 100,000 population across all younger age groups in both time periods.

*Trends in CVD Mortality Burden in Vietnam from 1990 to 2023*

More specifically, the ASMR decreased significantly from 1990 to 2000 before plateauing between 2000 and 2009 (APC: +0.15%) then subsequently resuming its decline till the next plateau from 2021 to 2023. The DALY rate followed a parallel trajectory, with an overall AAPC of -1.19% (95% CI: -1.23 to -1.15; p < 0.001), again with a significantly steeper decline in females (AAPC: -1.33%, 95% CI: -1.36 to -1.30) than in males (AAPC: -1.11%, 95% CI: -1.17 to -1.08). Moreover, prevalence trends were markedly divergent by sex. While female ASPR declined significantly over the period (AAPC: -0.05%, 95% CI: -0.06 to -0.04), male prevalence increased significantly (AAPC: +0.11%, 95% CI: +0.09 to +0.12). The overall population prevalence showed a marginal net increase (AAPC: +0.03%, 95% CI: +0.02 to +0.03). This male-driven divergence culminated in a sharp surge from 2021 to 2023 (APC: +1.43%).

*The Burden of Heart Failure related to CVD in 2023*

Age-specific distributions revealed two distinct crossover patterns: in absolute case counts, females exceeded males from the 65–69 age group onwards (33,620 vs. 31,110), reflecting greater female longevity at older ages; in ASPR, males maintained higher rates than females through the 85–89 group (7,578.87 vs. 7,352.89 per 100,000), with females only surpassing males at age 90–94 (8,906.67 vs. 8,762.61) and 95 years and above (9,963.31 vs. 9,580.26).

*Projection of CVD Burden in Vietnam to 2050*

Both ARIMA (0,1,2) and ARIMA (0,1,1) models maintained robust performance during the COVID-19 robustness validation period (2020–2023). For age-standardized death rates, ARIMA(0,1,2) achieved RMSE of 4.15 per 100,000 (MAPE: 1.70%) with all four observations falling within 95% prediction intervals (coverage: 4/4), indicating well-calibrated uncertainty and model resilience to pandemic disruption. For absolute deaths, ARIMA(0,1,1) achieved RMSE of 4,180 deaths (MAPE: 1.61%) with 75% coverage (3/4 observations within 95% CI); the 2021 observation fell outside the prediction interval, likely reflecting COVID-19-related excess mortality fluctuations (Figure S6A-B).

### Supplemental discussion

Sex disparities in CVD mortality were substantial, with male ASMR approximately twice that of females, reflecting a convergence of higher behavioral risk exposure and greater hypertension burden. Vietnam's striking sex disparity in smoking prevalence (approximately 45% in men versus 1–2% in women) and greater alcohol consumption are established drivers of stroke, IHD, and cardiomyopathy mortality.^4^ Additionally, males carry a double burden of higher hypertension prevalence and markedly lower blood pressure control rates — only 7.8% versus 16.7% in females — directly amplifying their CVD risk.^5^ In contrast, RHD mortality was higher in females, while absolute HF cases exceeded those of males from age ≥65 — reflecting greater female longevity and the steep age-dependence of HF risk. These findings underscore that sex-specific cardiovascular prevention strategies must account for subtype heterogeneity rather than treating male predominance as a universal pattern.

Vietnam's CVD burden between 1990 and 2023 reflects a nation at a critical epidemiological crossroads — achieving meaningful reductions in age-specific mortality while confronting an absolute burden that clinical progress alone cannot contain. The country's stroke-dominant profile, substantial sex disparities, and rapidly shifting metabolic risk landscape collectively define a CVD fingerprint demanding targeted, context-specific responses. Projections to 2050 make the urgency unmistakable: without a fundamental reorientation toward aging-specific prevention and chronic cardiovascular care, Vietnam risks being overwhelmed by the demographic tide of its own success. These findings provide the most comprehensive national evidence base to date for Vietnamese policymakers and the broader ASEAN cardiovascular community to act decisively — and equitably — before that window closes.

### Table S1. Average Annual Percent Change (AAPC) of Age-Standardized CVD Burden Metrics in Vietnam, 1990–2023

| Metric/ Population | AAPC (%) | 95% CI | P-value |
| --- | --- | --- | --- |
| Prevalence | | | |
| Both | 0.0293* | (0.0245 to 0.0338) | < 0.001 |
| Female | -0.0510* | (-0.0576 to -0.0448) | < 0.001 |
| Male | 0.1060* | (0.0926 to 0.1168) | < 0.001 |
| Incidence | | | |
| Both | -0.3299* | (-0.3458 to -0.3113) | < 0.001 |
| Female | -0.4243* | (-0.4381 to -0.4088) | < 0.001 |
| Male | -0.2575* | (-0.2709 to -0.2423) | < 0.001 |
| Deaths | | | |
| Both | -1.3435* | (-1.4062 to -1.3075) | < 0.001 |
| Female | -1.5870* | (-1.6250 to -1.5512) | < 0.001 |
| Male | -1.2101* | (-1.2547 to -1.1799) | < 0.001 |
| DALYs | | | |
| Both | -1.1940* | (-1.2336 to -1.1521) | < 0.001 |
| Female | -1.3281* | (-1.3622 to -1.2982) | < 0.001 |
| Male | -1.1113* | (-1.1707 to -1.0771) | < 0.001 |

* Indicates that the AAPC is significantly different from zero at the alpha = 0.05 level.

### Table S2. Model Validation Performance (2010-2019)

| Model | ASMR | | | | Deaths | | | |
| --- | --- | --- | --- | --- | --- | --- | --- | --- |
|  | RMSE | MAPE | AIC | LB p value | RMSE | MAPE | AIC | LB p value |
| ARIMA (0,1,2)* | 3.40 | 1.38% | 126.84 | 0.214 | 2,243 | 1.06% | 505.66 | 0.430 |
| ARIMA (0,1,1)† | 3.52 | 1.43% | 128.71 | 0.012 | 1,992 | 0.92% | 506.19 | 0.094 |
| Poisson (linear) | 7.31 | 2.85% | - | - | 3,245 | 1.48% | - | - |
| ARIMA (1,1,2) | 9.49 | 3.86% | 121.71 | 0.939 | 7,042 | 3.41% | 502.56 | 0.974 |
| ARIMA (1,1,1) | 10.85 | 4.38% | 119.76 | 0.940 | 8,158 | 3.91% | 500.56 | 0.973 |
| ARIMA (1,1,0) | 11.43 | 4.60% | 118.24 | 0.884 | 8,801 | 4.18% | 499.27 | 0.921 |
| Auto ARIMA | 15.96 | 6.26% | 113.20 | 0.701 | 11,270 | 5.22% | 481.14 | 0.971 |
| ETS | 15.97 | 6.27% | 142.12 | 0.952 | 11,269 | 5.22% | 530.40 | 0.979 |
| Poisson (quadratic) | 52.91 | 20.27% | - | - | 54,313 | 23.52% | - | - |

ASMR = age-standardized mortality rate; RMSE = root mean square error; MAPE = mean absolute percentage error; AIC = Akaike Information Criterion; LB = Ljung-Box test at lag 6.

*Selected final model for ASMR.

†Selected final model for absolute deaths.

All models trained on 1990–2009 data.

### Figure S1. Age-specific rates of cardiovascular disease burden by sex in Vietnam, 2023

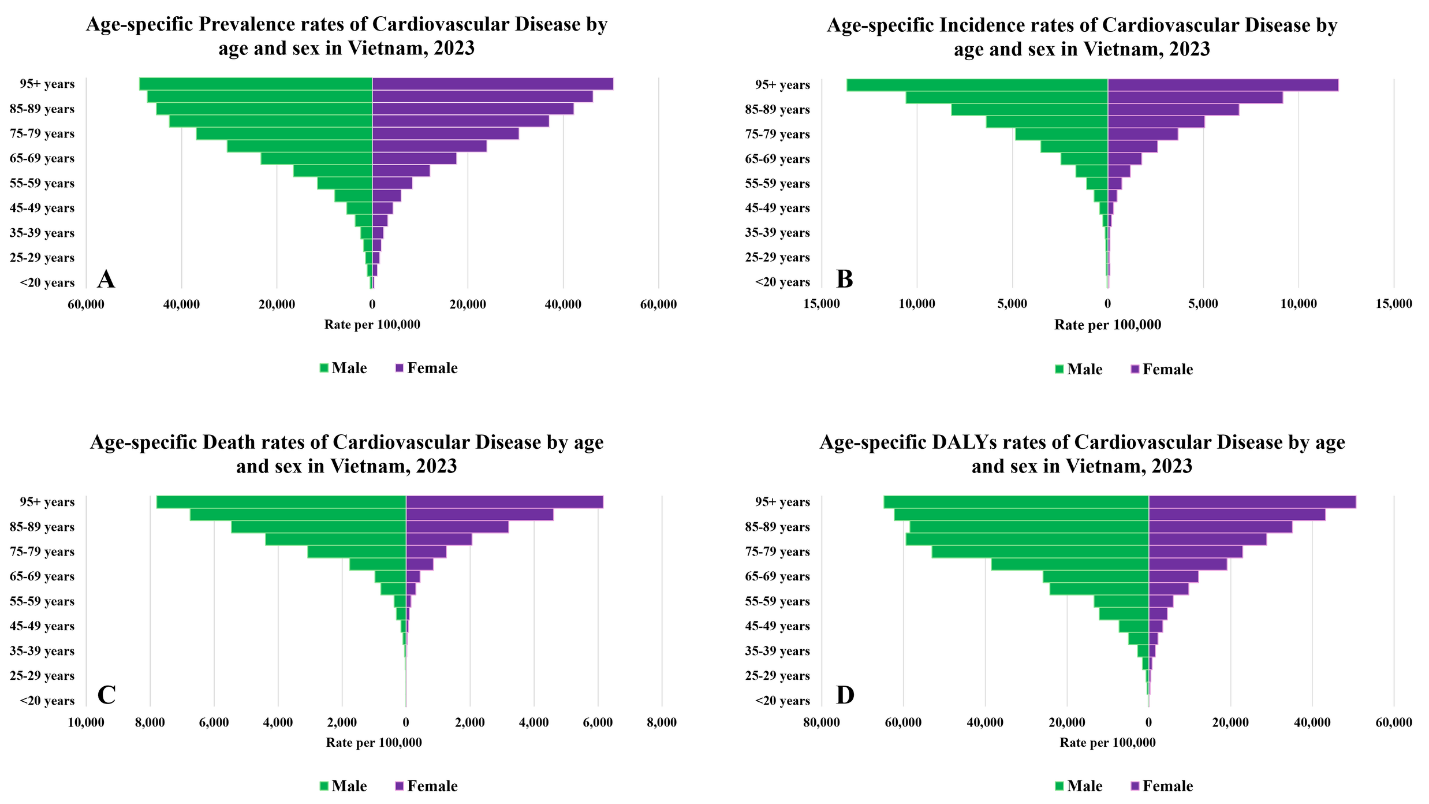

### Figure S2. Decomposition of changes in cardiovascular disease mortality burden in Vietnam, 1990–2023

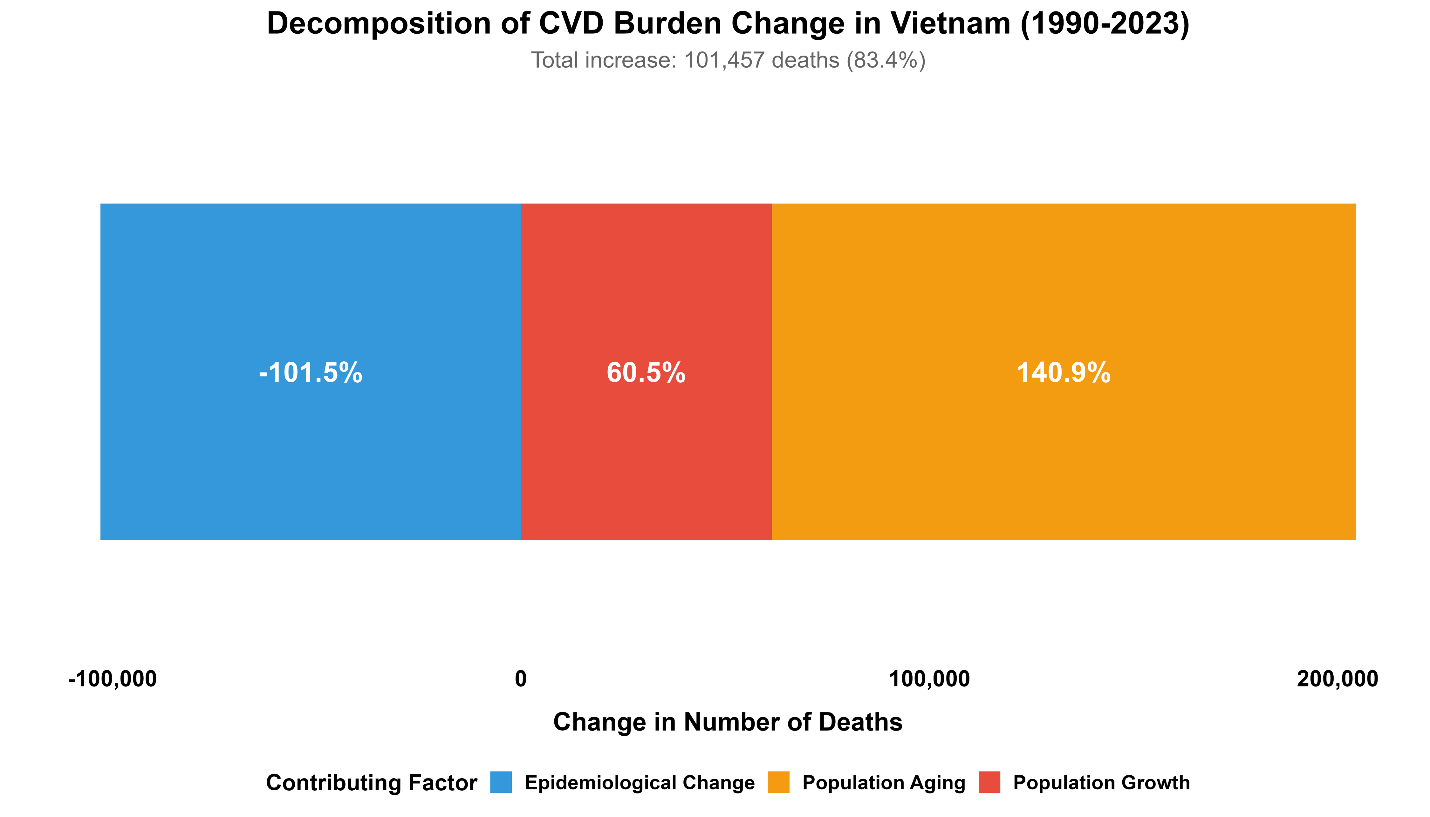

### Figure S3. Prevalent cases and prevalence rates of heart failure due to CVD in male and female across different age groups in Vietnam in 2023

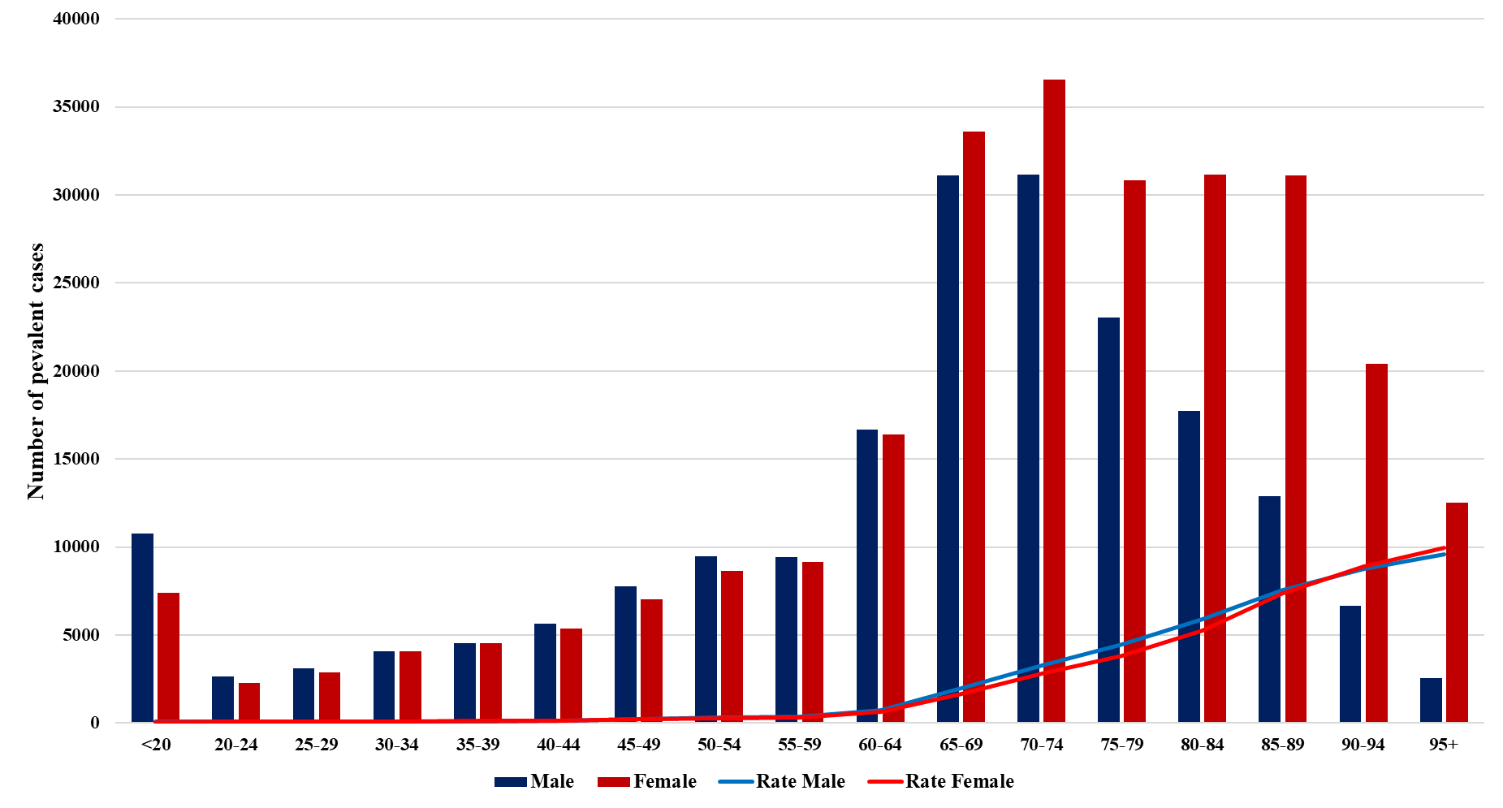

### Figure S4. Age-standardized cardiovascular disease DALYs rates attributable to cardiovascular risk factors in Vietnam, 1990 and 2023

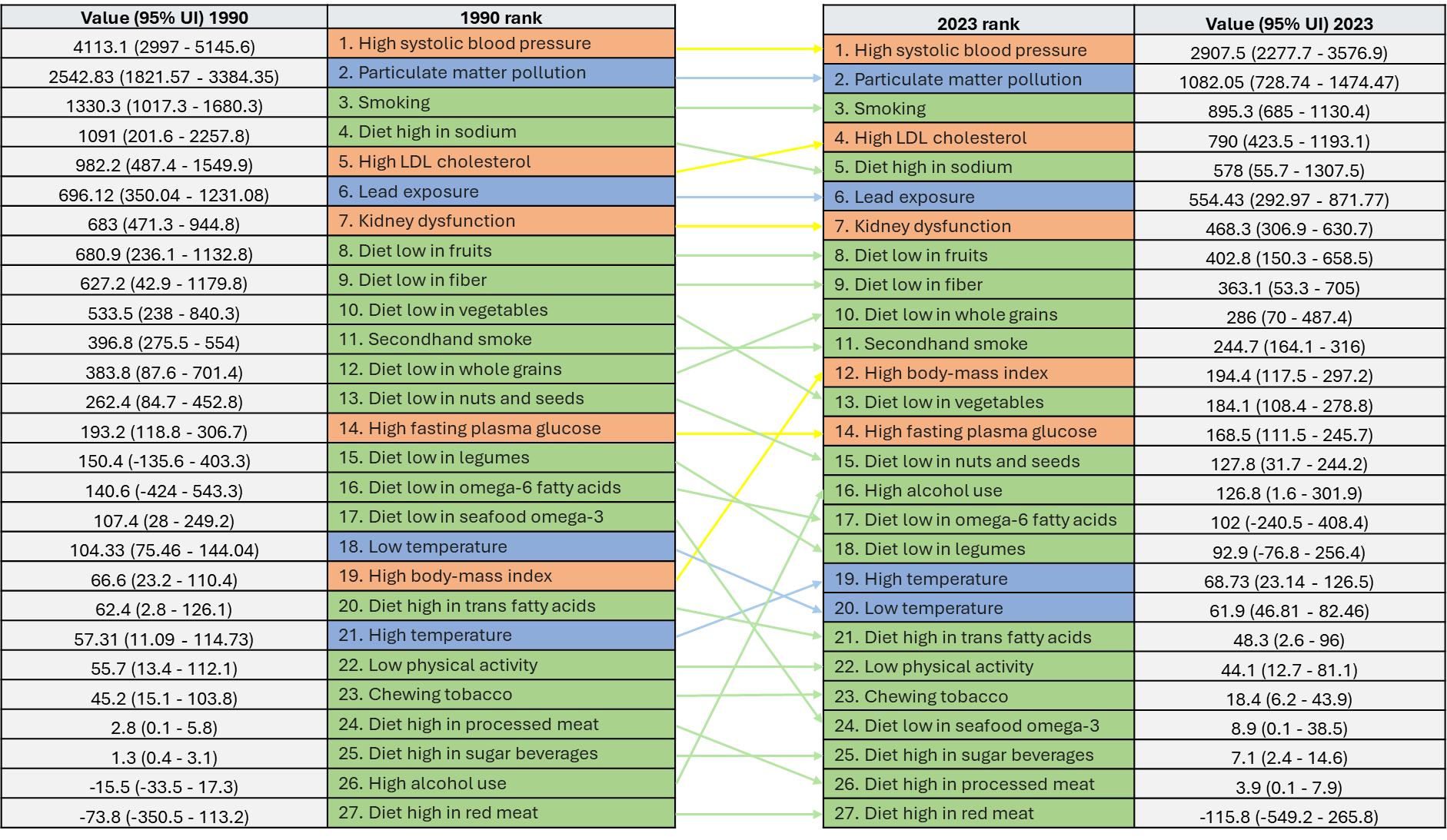

### Figure S5. Comparison of performance between nine models for forecasting age-standardized death rate and absolute deaths number through 2050

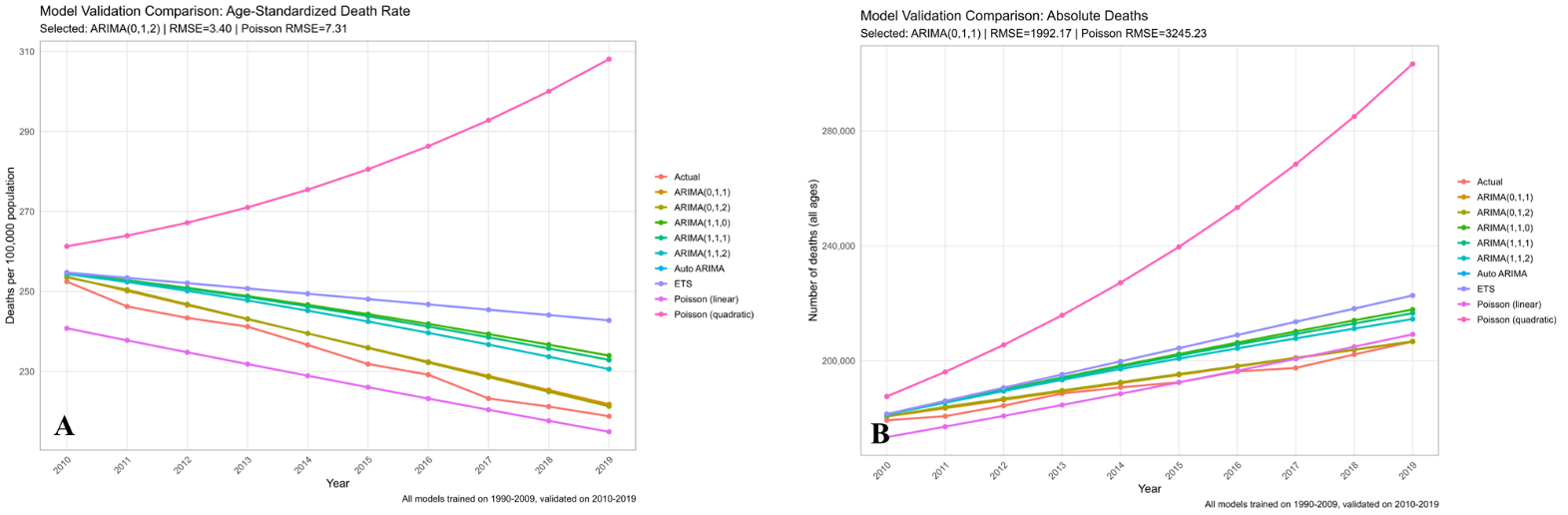

### Figure S6. COVID-19 period validation of ARIMA projection models for cardiovascular disease mortality in Vietnam, 2020–2023

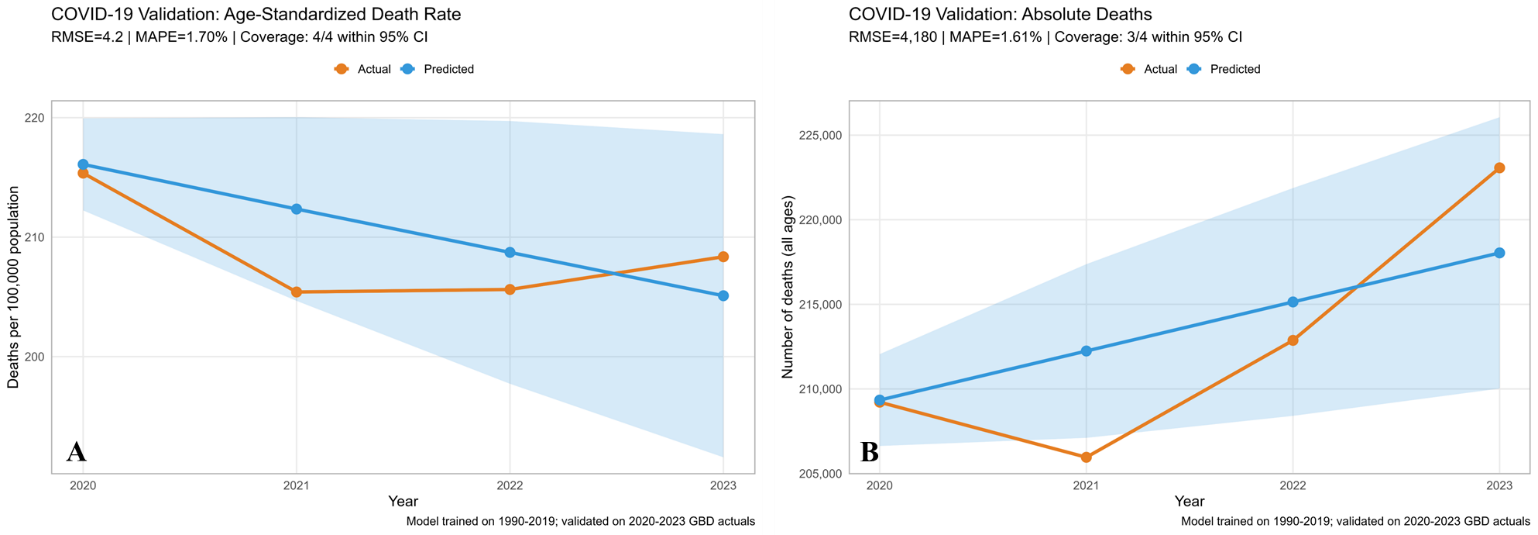
